# Deep learning enabled analysis of cardiac sphericity

**DOI:** 10.1101/2022.07.20.22277861

**Authors:** Milos Vukadinovic, Alan C. Kwan, Victoria Yuan, Michael Salerno, Daniel C. Lee, Christine M. Albert, Susan Cheng, Debiao Li, David Ouyang, Shoa L. Clarke

## Abstract

Quantification of chamber size and systolic function is a fundamental component of cardiac imaging, as these measurements provide a basis for establishing both diagnosis and appropriate treatment for a spectrum of cardiomyopathies. However, the human heart is a complex structure with significant uncharacterized phenotypic variation beyond traditional metrics of size and function. Characterizing variation in cardiac shape and morphology can add to our ability to understand and classify cardiovascular risk and pathophysiology. We describe deep learning enabled measurement of left ventricle (LV) sphericity using cardiac magnetic resonance imaging data from the UK Biobank and show that among adults with normal LV volumes and systolic function, increased sphericity is associated with increased risk for incident atrial fibrillation (HR 1.31 per SD, 95% CI 1.23-1.38), cardiomyopathy (HR 1.62 per SD, 95% CI 1.29-2.02), and heart failure (HR 1.24, 95% CI 1.11-1.39), independent of traditional risk factors including age, sex, hypertension, and body mass index. Using genome-wide association studies, we identify four loci associated with sphericity at genome-wide significance. These loci harbor known and suspected cardiomyopathy genes. Through genetic correlation and Mendelian randomization, we provide evidence that sphericity may represent a subclinical manifestation of non-ischemic cardiomyopathy.

## Introduction

The heart is a complex three-dimensional structure exposed to continuous dynamic stress. Genetic risk factors, environmental stressors, and age-associated changes impact cardiac morphology and function over the lifespan in a multitude of ways. Dilation of cardiac chambers and/or decline in systolic function are key indicators of disease, and conventional imaging assessments aim to quantify such changes. In turn, there has been significant interest in understanding the genetic basis for variation in such phenotypes^1,2^.

The study of cardiac morphology may provide additional clinical and genetic insights. Specifically, large biobanks with cardiac imaging data now offer an opportunity to define and analyze variation in cardiac morphology that is incompletely quantified by traditional measurements but present in normal populations^3^. However, precise phenotyping using imaging data is challenged by variability in image acquisition and human measurement^4^. Further, manual phenotyping is time-consuming and not feasible for testing novel phenotypes at larger scale. These challenges can be addressed by recent advances in deep learning and computer vision which allow for high throughput automated measurements of cardiac structures^5–8^, and may identify previously uncharacterized clinically-relevant phenotypic variation^3,9,10^.

Cardiomyopathies of different etiologies often result in a similar end-stage phenotype of a more round, spherical ventricle. Among patients with cardiac disease, left ventricle (LV) sphericity index has been previously associated with adverse outcomes and progression of disease, most commonly in echocardiography studies^11–16^. We hypothesized that within the spectrum of normal LV chamber size and systolic function, there exists variation in LV sphericity, and this variation may be a marker of cardiac risk with genetic underpinnings. To test this hypothesis, we applied automated deep-learning segmentation to cardiac magnetic resonance imaging (MRI) data available in the UK Biobank. We show that among patients with normal LV chamber size and systolic function, the sphericity index predicts incident cardiovascular diseases, including cardiomyopathy, atrial fibrillation (AF), and heart failure. Genetic analysis suggests a shared architecture between sphericity and non-ischemic cardiomyopathy (NICM), with NICM as a possible causal factor for LV sphericity among individuals with normal LV size and function.

## Results

### Measurement of the sphericity index using a convolutional neural network

In order to examine the clinical relevance and genetic influences of LV shape in the setting of normal size and function, we defined our main study cohort (n = 38,897) as UK Biobank participants with cardiac MRIs demonstrating normal LV end-diastolic volume, normal LV end-systolic volume, and normal LV ejection fraction (**Supplementary Figure 1**). **Table 1** shows the study cohort baseline characteristics. We applied a fully convolutional neural network for semantic segmentation of cardiac chambers to cardiac MRIs from the UK Biobank. During end-diastole in each segmented a 4-chamber image, we defined the smallest rectangle that fully encompassed the LV blood pool and measured the LV long axis length and short axis length. We defined the sphericity index as the ratio of the short axis length to the long axis length (**Figure 1a**). **Figure 1b** provides representative examples of the observed sphericity index in our study cohort. Sphericity index was similarly normally distributed in both men and women (**Figure 1c**) and showed a slight trend of increasing with age (**Figure 1d**).

**Table 1.** Cohort baseline characteristics.

| **Characteristic** | **Mean or n** |
| --- | --- |
| N | 38897 |
| Age at MRI | 54.9 ± 7.6 |
| Male | 17983 (46.2%) |
| Self-identified White British | 32965 (84.7%) |
| Body mass index (kg/m<sup>2</sup>) | 26.5 ± 4.2 |
| Hypertension | 2388 (6.1%) |
| Pulse rate | 67.7 ± 10.7 |
| LV ejection fraction (%) | 57.0 (4.47) |
| LV end diastolic volume (mL) | 135 ± 26.4 |
| LV end systolic volume (mL) | 58.4 ± 13.4 |

**Figure 1.**
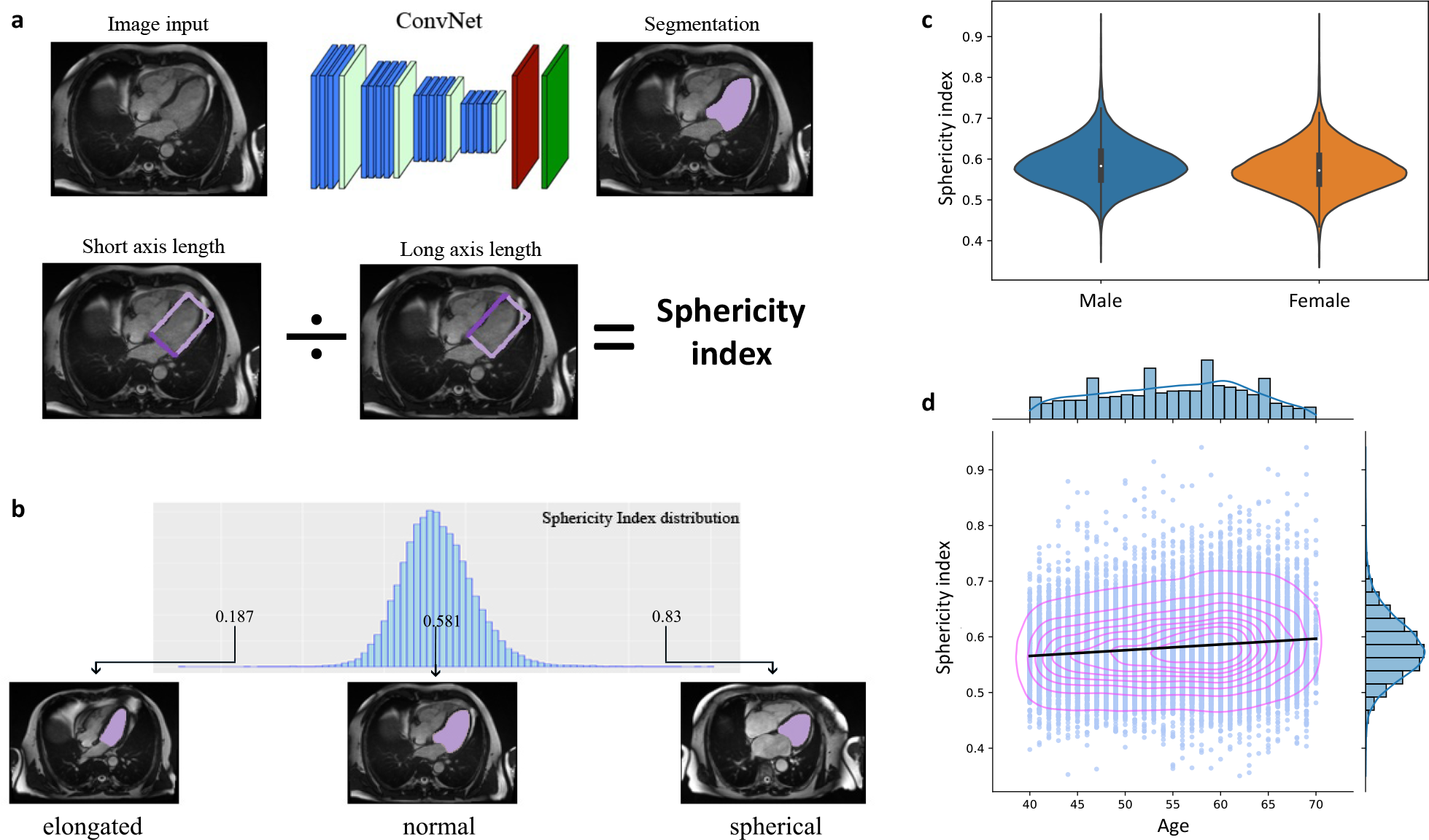
Deep learning enabled measurement of left ventricular sphericity index. **a**, Automated image segmentation of cardiac chambers was performed by applying a fully convolutional neural net to 4-chamber view cardiac MRI images. Sphericity index was defined by the ratio of the short and long axis lengths of a bounding box of the left ventricle. **b**, Distribution of the sphericity index in the study population and representative examples. **c**, The sphericity index is similarly normally distributed in male and female subjects. **d**, The sphericity index showed a small positive correlation with age. Concentric contour lines reflect increasing density of data point.

### Phenome-wide association study of left ventricular sphericity

We broadly assessed for clinical associations with LV sphericity using a phenome-wide association study (PheWAS) approach (**Figure 2**), and we identified AF as the top association by P value (Pearson *r* 0.05, P 9.5×10^−14^). In comparison, LV long axis length had no association with AF (*r* -0.007, P 0.3), while LV short axis length showed a similar association with AF (*r* 0.05, P 2.6×10^−14^). Notably however, AF only ranked 8^th^ by P value for short axis length. Other associations, including pulse rate (*r* -0.15, P 2.4×10^−129^), systolic blood pressure (*r* 0.09, P 2.3×10^−43^), and obesity (*r* 0.08, P 3.2×10^−38^) were substantially stronger for PheWAS of LV short axis length (**Supplementary Figure 2**). These results show that while metrics of shape may have overlapping clinical associations with metrics of size, analyses of shape may help highlight some associations by attenuating non-specific associations related to anthropometrics.

**Figure 2.**
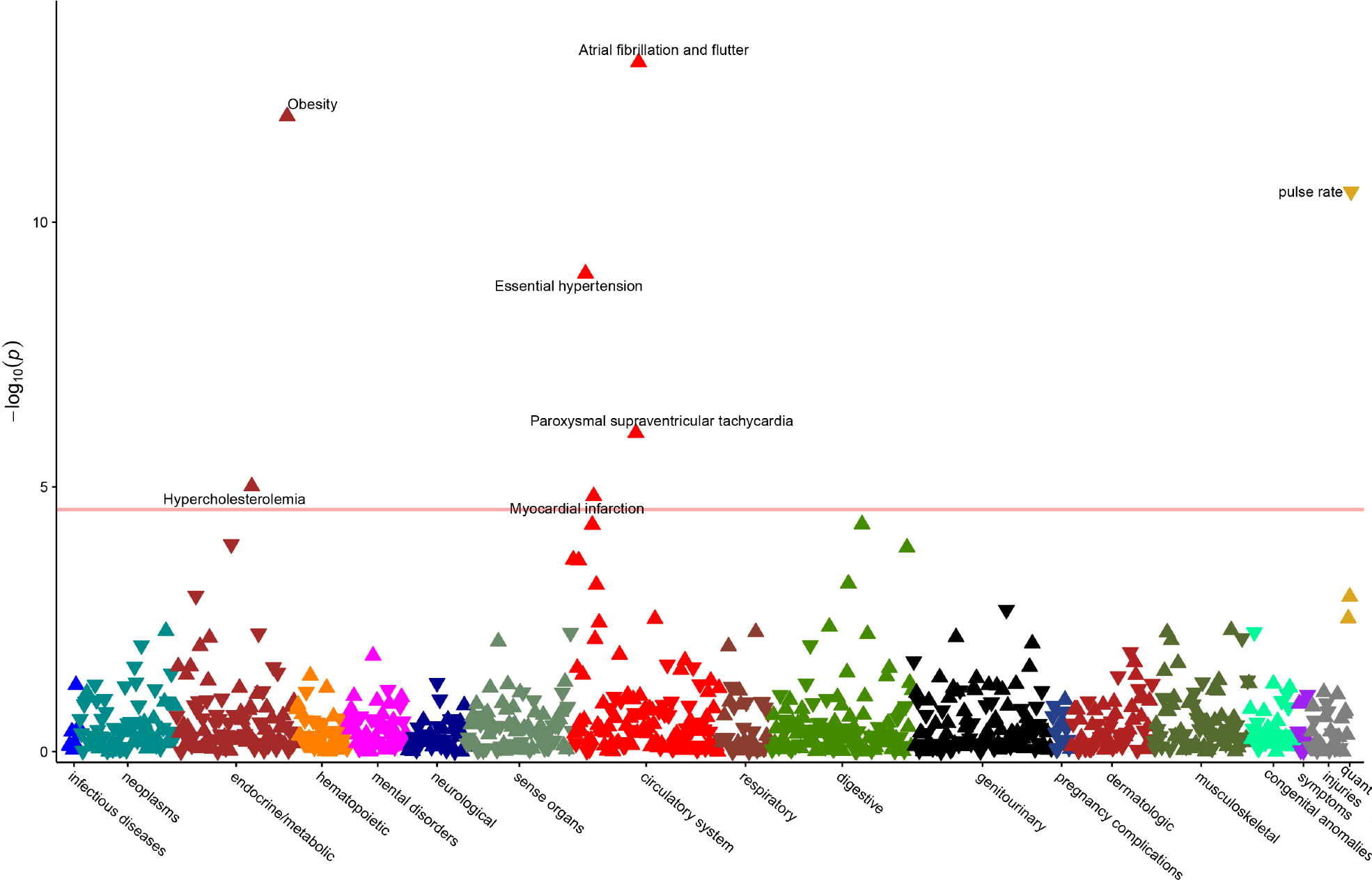
Phenome-wide association study of left ventricular sphericity index. Upward pointing triangles represent positive correlations with sphericity index, and downward pointing triangles represent negative correlations. The horizontal red line reflects the threshold of Bonferroni significance for 966 tests (5×10^−5^). Associations meeting Bonferroni significance are labeled.

### Left ventricular sphericity predicts incident disease

We next assessed whether the sphericity index predicts incident disease. Given prior studies^12–17^ and the results of our PheWAS analysis, we chose four outcomes available through first-occurrence data from the UK Biobank: AF, cardiomyopathy, heart failure, and cardiac arrest. We performed Cox analysis of incident outcomes with adjustments for age at MRI, sex, BMI, pulse rate, and baseline hypertension. We found that the sphericity index predicted incident AF, cardiomyopathy, and heart failure but not cardiac arrest (**Figure 3**). The hazard ratios (HR) associated with a 1-SD increase in sphericity index were 1.31 (95% CI 1.23-1.38, P <2×10^−16^), 1.62 (95% CI 1.29-2.02, P 2.4×10^−5^), and 1.24 (95% CI 1.11-1.39, P 1.8×10^−4^) for AF, cardiomyopathy, and heart failure respectively.

**Figure 3.**
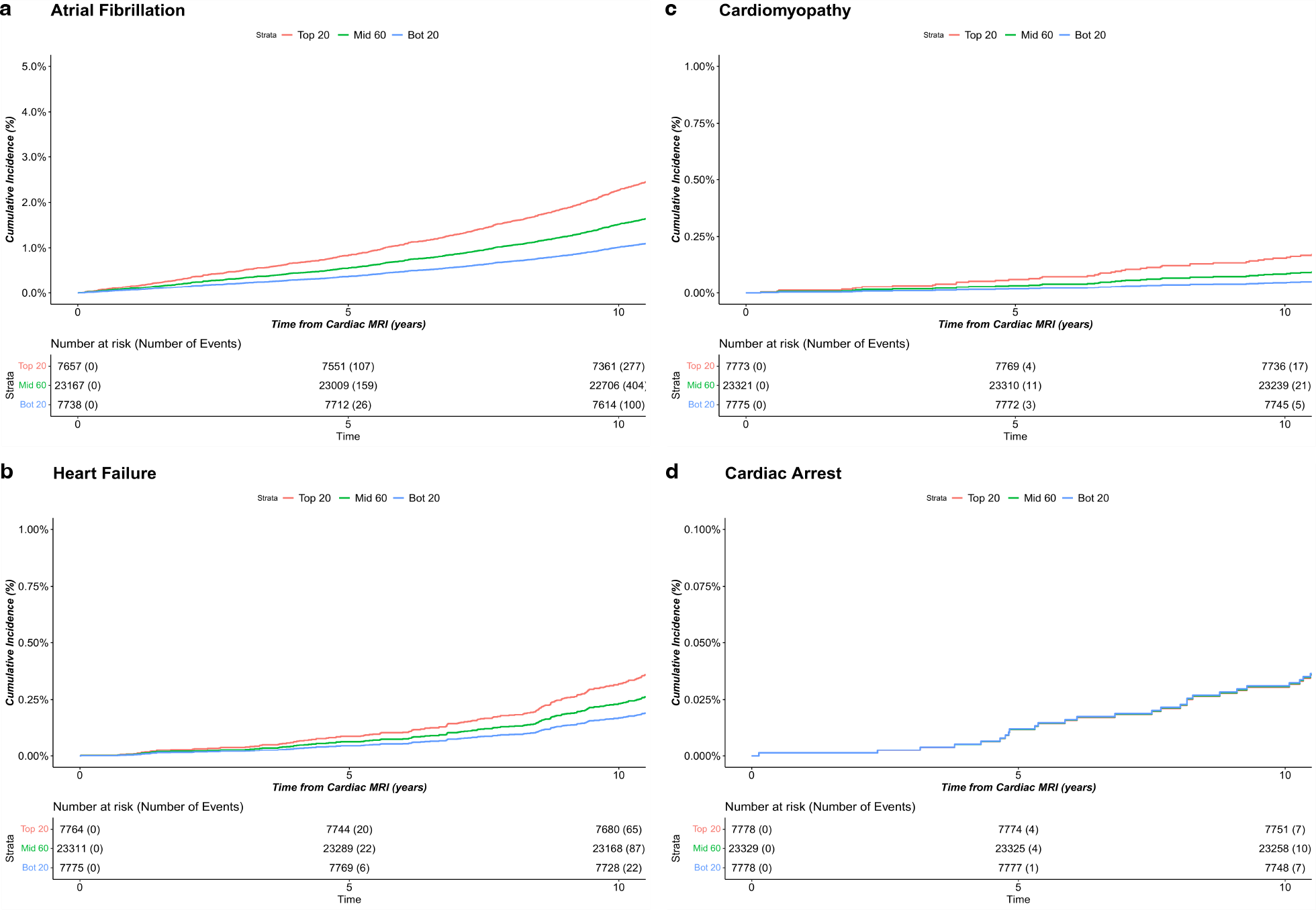
Risk of incident disease stratified by quintiles of sphericity index. Time-to-event analysis was performed from time of MRI to first occurrence of (**a**) atrial fibrillation, (**b**) cardiomyopathy, (**c**) heart failure, and (**d**) cardiac arrest. Models were adjusted for age at MRI, sex, body mass index, hypertension, and pulse rate. Plots show cumulative incidence of disease stratified by sphericity index quintiles, comparing the top 20%, middle 60%, and bottom 20%.

### Genome-wide association study of left ventricular sphericity

We then performed a genome-wide association study (GWAS) of common genetic variants and LV sphericity index and compared the results to the GWAS of the LV long and short axis lengths (**Figure 4**). We identified four loci associated with LV sphericity index at genome-wide significance (P ≤ 5×10^−8^): *PLN, ANGPT1, PDZRN3, and HLA DR/DQ* (**Table 2**). *PLN* is gene well known for its causal role in dilated and arrhythmogenic cardiomyopathies^18,19^. *ANGPT1* has not been associated with cardiomyopathy to date, but mouse experiments have demonstrated a critical role in mammalian heart development^20,21^. The *PDZRN3* locus has been previously associated with electrocardiographic characteristics^22^ and LV fractal structure in prior GWAS^3^, and regulation of post-natal expression has been implicated in murine cardiac maturation and geometry^23^. The *HLA DR/DQ* locus has long been implicated in dilated cardiomyopathy^24,25^, and recent GWAS that included both normal and abnormal cardiac MRIs have identified this locus for both left and right chamber size metrics^2,26^.

**Table 2.**
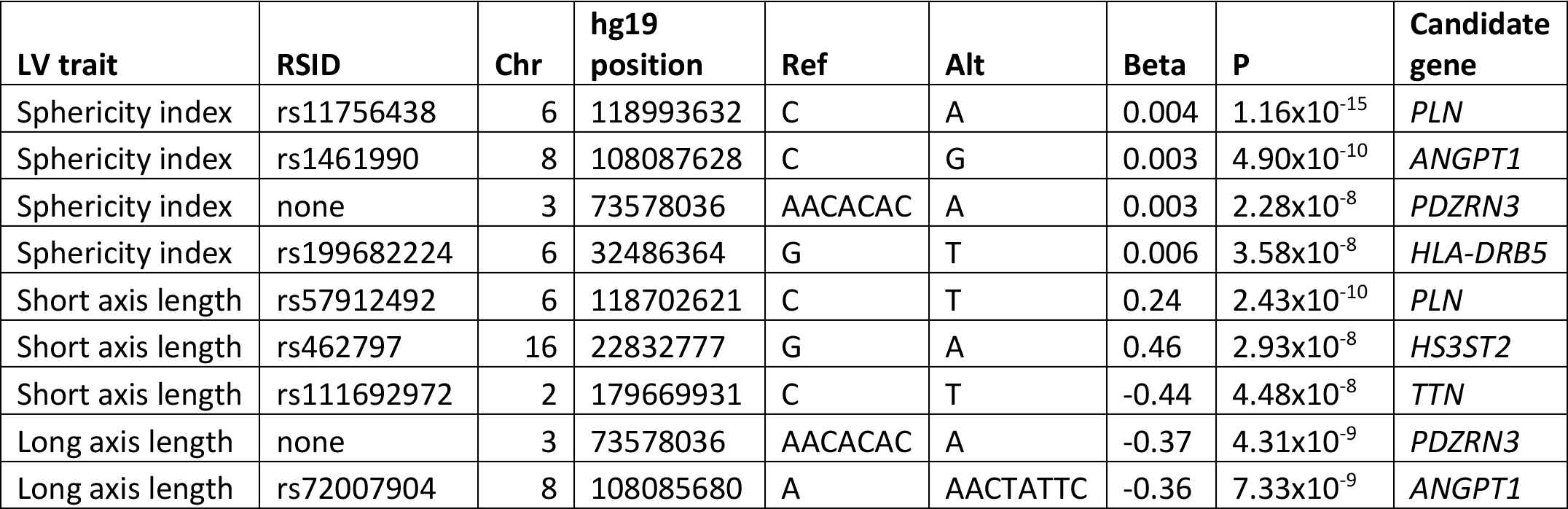
Lead variants and candidate genes for genome-wide significant loci of each left ventricle (LV) trait.

**Figure 4.**
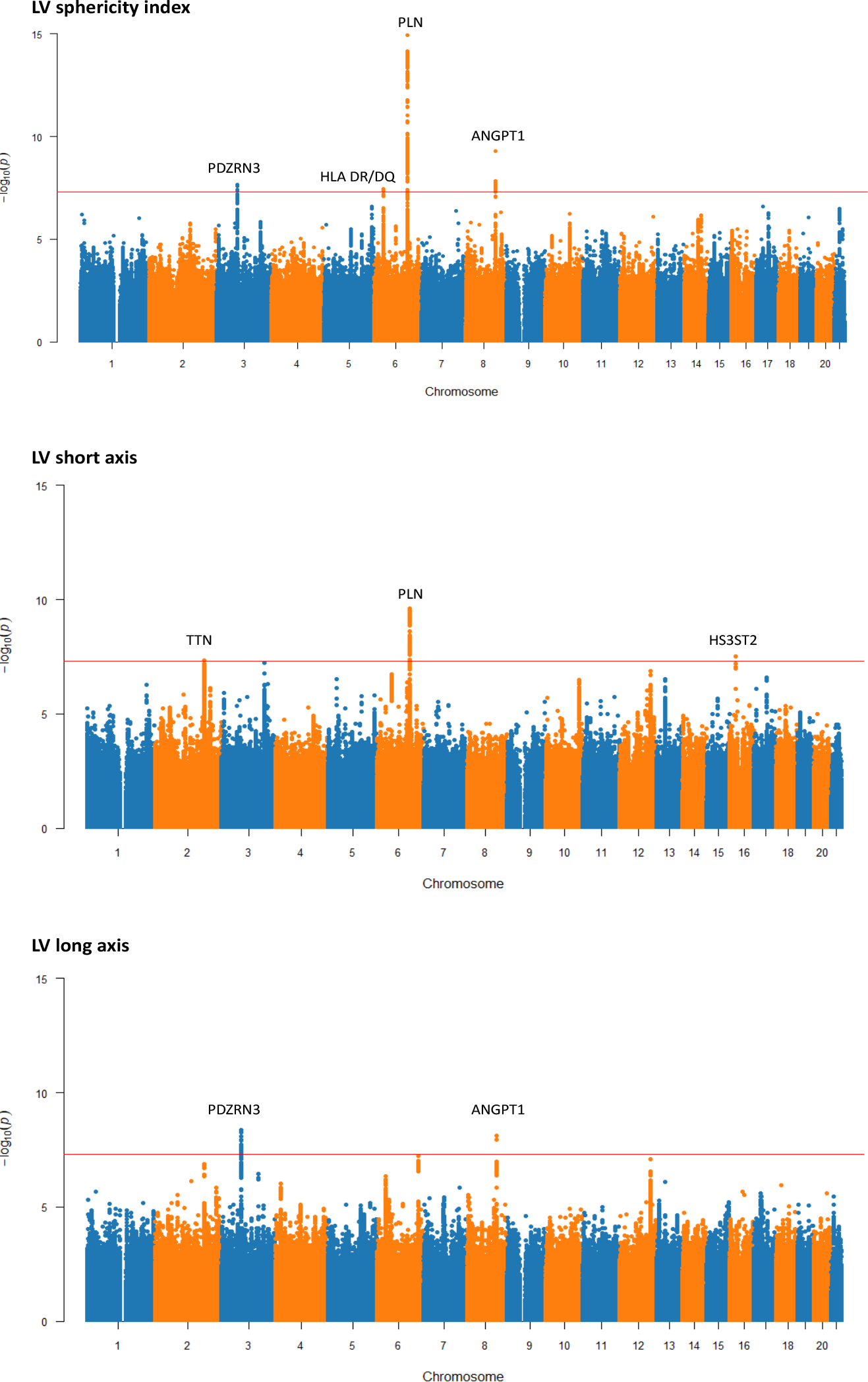
Manhattan plots for genome-wide associations studies of left ventricular sphericity index, short axis length, and long axis length. The red horizontal line represents a genome-wide significant P value of 5×10^−8^. Regions that reach genome-wide significant are labeled with candidate loci.

Comparing the GWAS of sphericity to LV short and long axis lengths highlights the value of studying shape-related phenotypes. Given the inherent interplay between sphericity and axis lengths, we expected to see overlap in genetic associations. Indeed, the genetic correlations with sphericity index were 0.64 (P 2.7×10^−25^) and -0.63 (P 6.8×10^−27^) for short axis and long axis lengths respectively. In terms of genome-wide significant loci, *PLN* was identified by both sphericity and short axis length but not long axis length. Notably, the significance of the association was substantially stronger for sphericity. Conversely, *ANGPT1* and *PDRZRN3* were identified by long axis length but not short axis length. Interestingly, the *TTN* gene was significant for short axis length (4.5×10^−8^) and suggestive for long axis length (P 1.5×10^−7^), but the association with sphericity was much weaker (P 2.1×10^−5^). This pattern might suggest that *TTN* impacts overall heart size more so than shape.

### Relationship between sphericity and cardiomyopathy

We explored the relationship between LV sphericity and non-ischemic cardiomyopathy (NICM) by first measuring the genetic correlation. Given the findings from our Cox analysis, we also assessed AF as a comparison. For each outcome, we used recent large GWAS^27,28^, and correlations were assessed by LD score regression, a technique that is robust to potential sample overlap. We found a significant correlation between sphericity index and cardiomyopathy (*rg* 0.42, P 0.01) but no genetic correlation between sphericity and AF (*rg* 0.04, P 0.5).

We then assessed for evidence of causal relationships using bidirectional two-sample Mendelian randomization by inverse variance weighted meta-analysis. To do so, we used summary statistics for NICM and AF from the FinnGen project^29^, a cohort independent of the UK Biobank. When assessing sphericity index as an exposure, we found no evidence for causality with either NICM or AF. We then assessed sphericity index as an outcome. When using AF as an exposure, we found no evidence for causality with sphericity (**Supplementary Table**). However, when using NICM as an exposure, we found a significant causal relationship for sphericity (beta 0.002, P 0.03). In sensitivity analyses, the relationship remained significant by weighted medians (beta 0.001, P 0.049) but not by MR Egger (beta 0.0007, P 0.6). Together, these findings suggest that cardiomyopathy may be causal for increased sphericity even among individuals with normal measurements of LV systolic function and volumes.

## Discussion

We have shown that the LV sphericity index predicts risk for incident AF, cardiomyopathy, and heart failure in healthy adults. Our genetic analyses suggest that LV sphericity has overlapping genetic architecture with clinical NICM, and the study of phenotypes related to organ shape may provide complimentary insight to the study of organ size. Overall, our data support the hypothesis that spherical LVs in healthy patients may reflect an early subclinical manifestation of cardiomyopathy.

We chose to study LV sphericity as a simple and interpretable metric of LV shape, motivated by clinical experience and prior studies that that suggest cardiac sphericity may be a marker of disease. Prior studies were limited by their focus on sphericity after the onset of clinical disease^11–16^, highlighting the relationship between shape and impaired contractility as spherical remodeling occurs secondary to disease or injury. We have added to these prior works by applying deep learning to conduct the first large-scale study of sphericity, while also assessing its clinical relevance and genetic architecture among subjects with normal LV chamber size and systolic function.

The main limitation of this study is the use of a single cohort. The UK Biobank is known to have a healthy cohort bias and does not represent a random sample of the UK population. Replication in additional cohorts will be necessary to validate the use of sphericity for clinical risk prediction. Additionally, in order to limit inflation due to population substructure, our GWAS included only a subset of unrelated participants of similar European ancestry. Lastly, sphericity index may not fully capture phenotypic variation within the left ventricle, as additional features of variable trabeculation, angle, and thickness simultaneously influence cardiac function.

In conclusion, this study demonstrates the utility of using deep learning and advanced imaging analysis to define and study non-traditional cardiac imaging risk biomarkers using the large-scale data that is increasingly available in biobanks. While conventional imaging metrics have significant diagnostic and prognostic value, some of these measurements have been adopted out of convenience or tradition. By representing a specific multi-dimensional remodeling phenotype, sphericity has emerged as a distinct morphologic trait with features not adequately captured by conventional measurements. We expect that the search space of potential imaging measurements is vast, and we have only begun to scratch at the surface of disease associations.

## Methods

### Cohort

The UK Biobank (UKB) is a population-based cohort that links genetic and phenotypic data for approximately 500,000 adult participants from the United Kingdom^30 31^. Our analyses focused on ∼48,000 participants who have undergone cardiac MRI^32^. A subset of this group that met quality control filters and that had normal MRI measurements (LV end-diastolic volume 88-218 mL, LV end-systolic volume 31-97 mL, and LV ejection fraction 48-70%)^33^ was used as the main cohort (**Supplementary Figure 1**). For the genome-wide association study (GWAS), we selected subjects from the main cohort who passed additional GWAS filters.

### Extraction of Sphericity Index Trait using Machine Learning

We downloaded cardiac MRI containing steady-state free precession image sequences of horizontal long-axis view. Each sequence is given as 210 × 208 × 50 matrix in DICOM format, with in-plane resolution 1.8 × 1.8 *mm*^2^. In the pre-processing phase, we converted DICOM images and manual annotations (.XML) to NIfTI format, which saves the sequence as a single 3d image for the sake of better file management.

We used a fully convolutional neural network^5,34^ for automated LV segmentation of MRI images. Trained weights were obtained from a prior work and inference was run on each video of each patient with cardiac MRIs in the UKB. For each individual, the semantic segmentation was manually evaluated and extreme outliers were excluded, as well as outliers in summary statistics provided from the UKB. We performed a segmentation quality check in two phases. We both heuristically checked (identifying when the segmentation was not continuous or implausibly small) and visually inspected segmentations to excluded patients with suboptimal segmentations.

For all images that passed quality check we created an automated bounding box around the segmentation of left ventricular blood pool (**Figure 1**a). The longer side of the rectangle was taken as LV long axis length (LVL), and the shorter side was taken as LV short axis length (LVS). Sphericity index is calculated as quotient of LVS and LVL 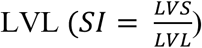. Higher value of sphericity index indicates more spherical appearance, and lower value indicates an elongated appearance of left ventricular chamber.

### PheWAS

Diagnostic data was downloaded as ICD10 codes^5^, processed, and saved as diagnostic groups organized by phecodes using a mapping file provided by PheWAS Catalog^7^. Phenome-wide association studies (PheWAS) ^35^ were performed by measuring the Pearson *r* correlation between sphericity index and each phecode. In addition, three quantitative measurements were assessed: pulse rate, systolic blood pressure, and diastolic blood pressure. A total of 966 correlations were assessed. Significant associations were identified as those with a P value below the Bonferroni-corrected threshold of 5×10^−5^.

### Cox Survival Analysis

We performed a time-to-event analysis to assess the association of sphericity index with incident atrial fibrillation, cardiomyopathy, heart failure, and cardiac arrest, using the UKB first occurrence data. These outcomes are provided by the UKB as part of a curated set of outcomes derived from health record diagnostic codes. We defined days without incident as the time from the first MRI visit to the first occurrence of the given outcome, death, or last follow-up (March 31st, 2021). For outcomes of atrial fibrillation, heart failure, and cardiomyopathy, participants with the prevalent disease at the time of the first MRI visit were excluded. We measured the association between the sphericity index and each outcome using a multivariable Cox proportional hazards model adjusted for age at MRI, sex, body mass index (BMI), pulse rate, and hypertension. For this analysis, the sphericity index was normalized to mean equaling 0 and standard deviation (SD) equaling 1, and the hazard ratio (HR) is reported as per 1-SD. We additionally stratified the cohort into the lower 20th, middle 60th, and upper 20th percent of the sphericity index and plotted adjusted cumulative incidence curves by strata.

### Genome-wide association study

We used the UKB imputed genotype calls in BGEN v1.2 format. Samples were genotyped using the UK BiLEVE or UK Biobank Axiom arrays. Imputation was performed using the Haplotype Reference Consortium panel and the UK10K+1000 Genomes panel ^30^. We used the QC files provided by UKB to create a GWAS cohort consisting of subjects who did not withdraw, were of inferred European ancestry, and were unrelated. Subjects with a genotype call rate < 0.98 were also removed. We considered variants with a minor allele frequency (MAF) ≥ 0.01, and we required genotyped variant to have a call rate ≥ 0.95 and imputed variants to have an INFO score ≥ 0.3. Variants with a Hardy-Weinberg equilibrium P value < 1×10^−20^ were excluded. GWAS was done on a Spark 3.1.1 cluster, using library Hail 0.2 with Python version 3.6. The GWAS as adjusted for age at MRI and sex. We used the conventional P value of 5×10^−8^ as the threshold for defining genome-wide significance.

### Genetic correlation

Genetic correlations were estimated using *ldsc version 1*.*01*^36^. For GWAS performed in this study (sphericity index, short axis length, long axis length), we used well imputed variants by filtering for INFO ≥ 0.9. For NICM and AF, we used summary statistics from previously published GWAS^27,28^, and we limited to well imputed variants by intersecting with HapMap3 variants.

### Mendelian randomization

Two-sample Mendelian randomization was conducted using the 2SampleMR version 0.5.6 R package. We used a bidirectional approach with LV sphericity index as the exposure for the outcomes of NICM and AF and with LV sphericity as an outcome with NICM and AF as exposures. We used FinnGen^29^ release 6 GWAS summary statistics for NICM (finngen_R6_I9_NONISCHCARDMYOP_STRICT) and AF (finngen_R6_I9_AF) to assure non-overlapping samples. The FinnGen NICM strict phenotype excludes hypertrophic cardiomyopathy. For each exposure, the instrument variable was created by identifying independent significant or suggestive single nucleotide polymorphisms (SNPs) that could be harmonized to the outcome of interest unambiguously. Independent SNPs were defined using a clumping distance of 10 megabases and an R^2^ threshold of 0.001. Both sphericity index and NICM had a limited number of genome-wide significant variants after excluding palindromic SNPs and indels, and we therefore included SNPs with a suggestive P value of <10^−6^. This resulted in an 11-SNP instrument variable for sphericity index and a 7-SNP instrument variable for NICM. For AF, we only considered genome-wide significant SNPs (≤5×10^−8^), resulting in a 34-SNP instrument variable. We used inverse-variance weighted meta-analysis as our primary approach, and we used median weighted meta-analysis and MR Egger as sensitivity analyses.

## Data Availability

UK Biobank data is available with an approved study protocol.

https://www.ukbiobank.ac.uk/

