## Supplemental Materials for "Deep learning enabled analysis of cardiac sphericity"

**Supplementary Table. Two-sample Mendelian randomization using inverse-variance weighted meta-analysis.**

| Exposure | Outcome | Beta | SE | P value |
| --- | --- | --- | --- | --- |
| SI | NICM | 3.3 | 6.5 | 0.6 |
| SI | AF | 1.6 | 3.7 | 0.7 |
| NICM | SI | 0.001 | 0.0008 | 0.03 |
| AF | SI | -0.0004 | 0.001 | 0.7 |
| SI = sphericity index; NICM = non-ischemic cardiomyopathy; AF = atrial fibrillation; SE = standard error | | | | |

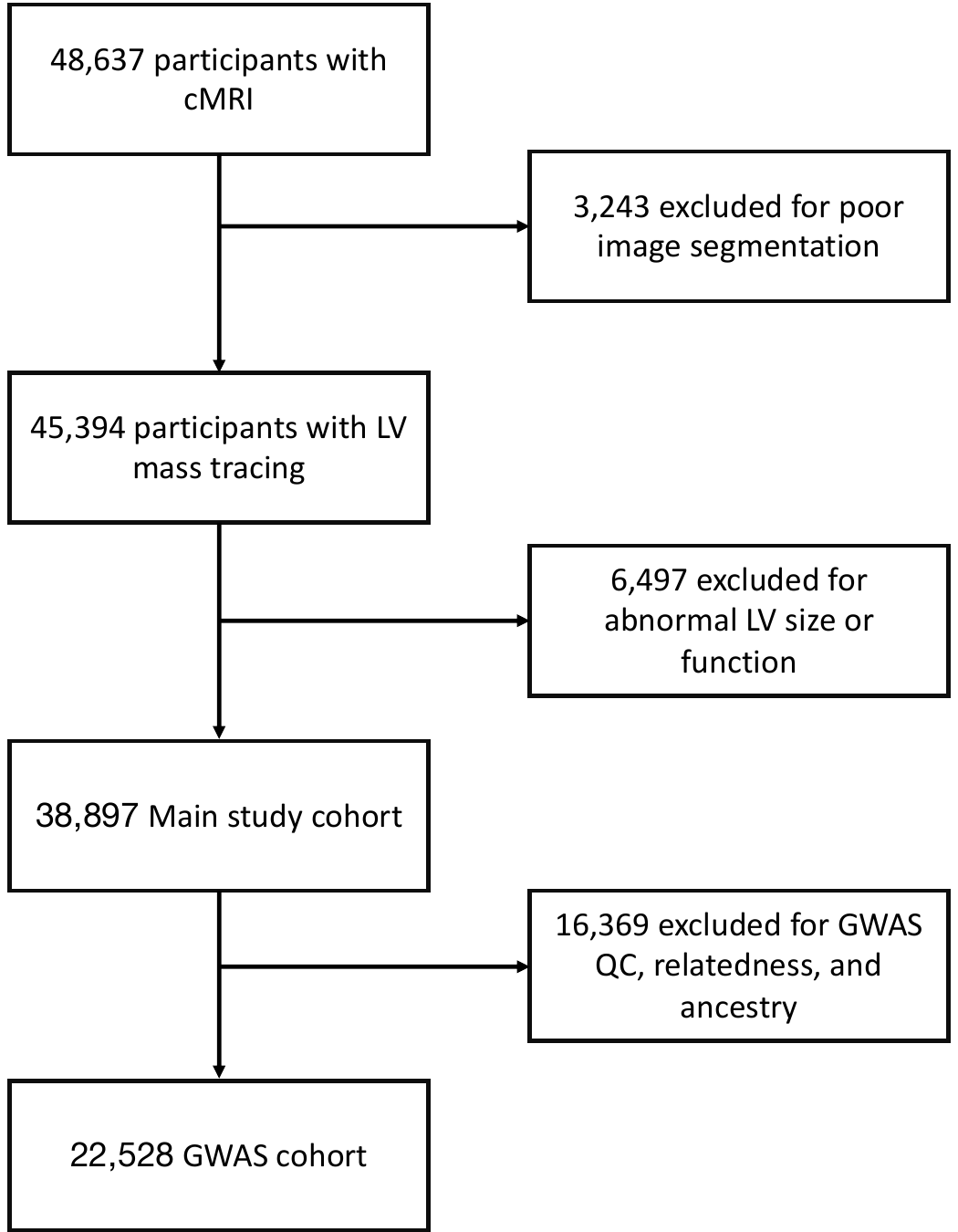

**Supplementary Figure 1. Cohort diagram**

| **a** | **b** |
| --- | --- |
| 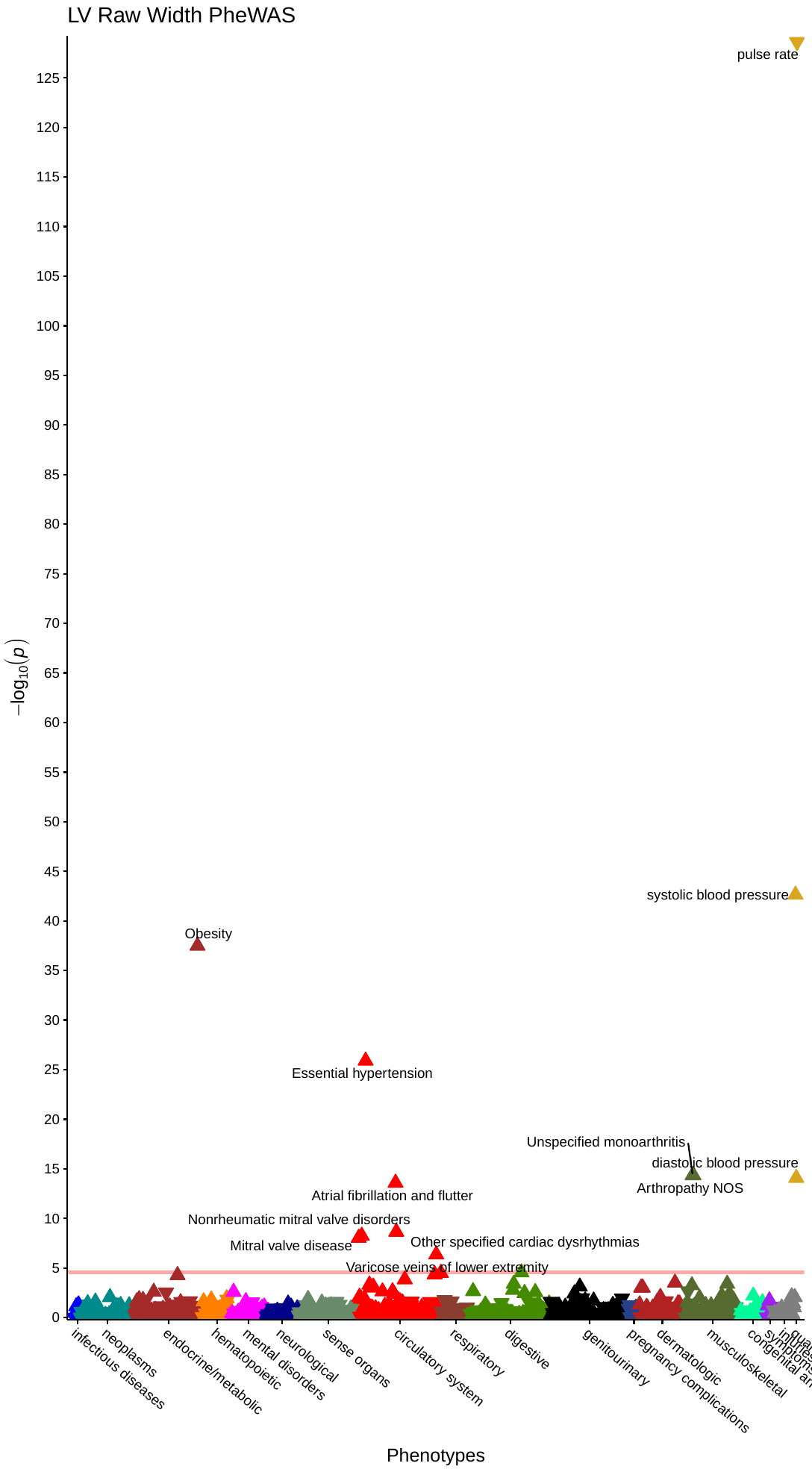 | 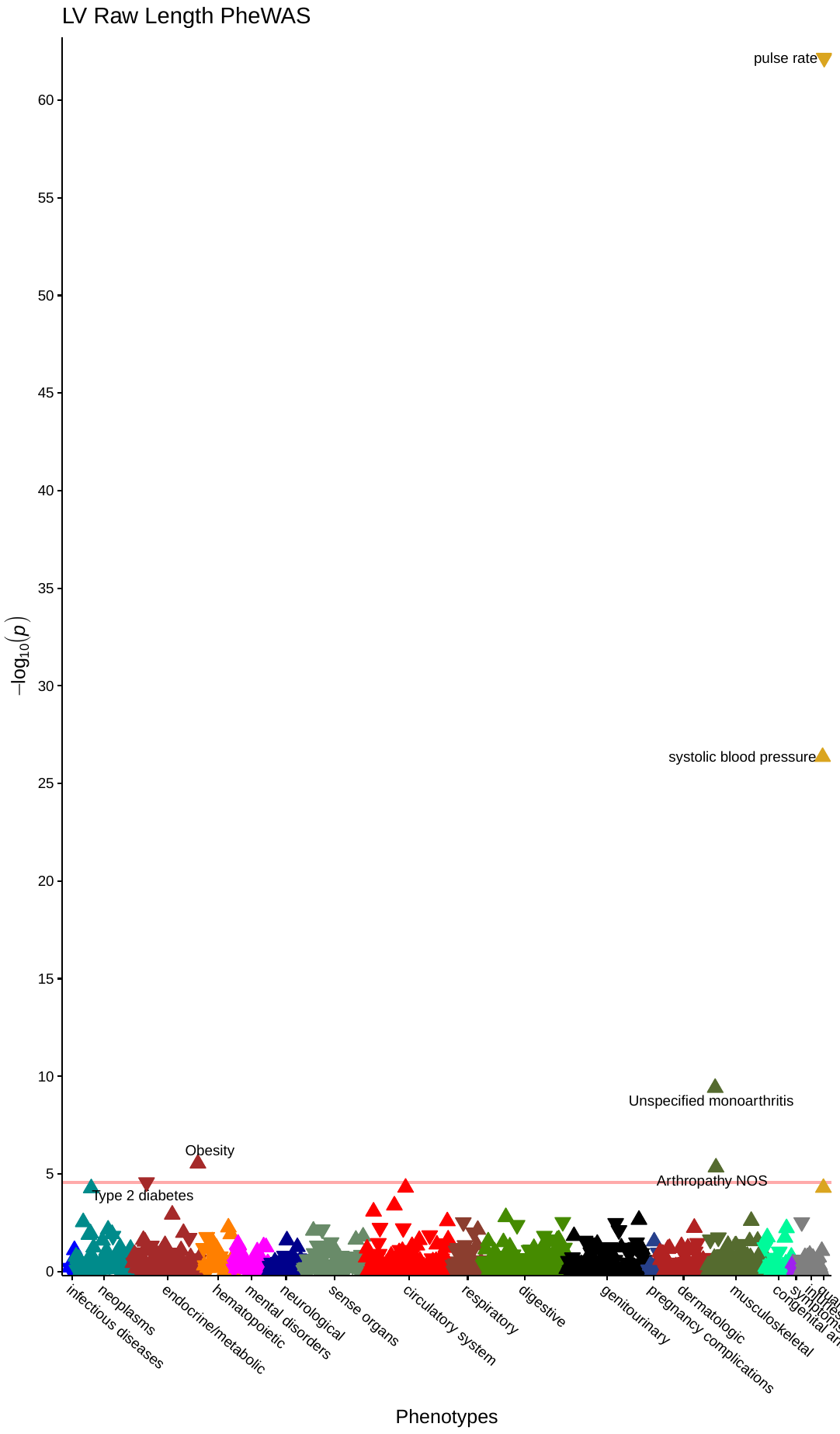 |

**Supplementary Figure 2. Phenome-wide association studies of (a) left ventricular short axis length and (b) long axis length.** Upward pointing triangles represent positive correlations with sphericity index, and downward pointing triangles represent negative correlations. The horizontal red line reflects the threshold of Bonferroni significance for 966 tests (5x10^-5^). Associations meeting Bonferroni significance are labeled.
